# Accounting for health inequities in the design of contact tracing interventions: a rapid review

**DOI:** 10.1101/2021.03.01.21252692

**Authors:** Isadora Mathevet, Katarina Ost, Lola Traverson, Kate Zinszer, Valéry Ridde

## Abstract

**Context:** Contact tracing has been a central COVID-19 transmission control measure. However, without the consideration of the needs of specific populations, public health interventions can exacerbate health inequities.

**Purpose:** The purpose of this rapid review was to determine if and how health inequities were included in the design of contact tracing interventions in epidemic settings.

**Method:** We conducted a search of the electronic databases MEDLINE and Web of Science. Our inclusion criteria included articles that: (i) described the design of contact tracing interventions, (ii) have been published between 2013 and 2020 in English, French, Spanish, Chinese, or Portuguese, (iii) and included at least 50% of empiricism, according to the Automated Classifier of Texts on Scientific Studies (ATCER) tool. We relied on various tools to extract data.

**Result:** Following the titles and abstracts screening of 230 articles, 39 articles met the inclusion criteria. Only seven references were retained after full text review. None of the selected studies considered health inequities in the design of contact tracing interventions.

**Conclusion:** The use of tools/concepts for incorporating health inequities, such as the REFLEX-ISS tool, and “proportionate universalism” when designing contact tracing interventions, would enable practitioners, decision makers, and researchers to better consider health inequities.

## Introduction

Contact tracing plays a key role in controlling communicable diseases by seeking to break the chain of transmission between individuals. It is recommended by the World Health Organization (WHO) as part of the global strategy for COVID-19, which also includes case identification, isolation, testing, care, and quarantine.^1^ Contact tracing consists of identifying and monitoring individuals who have been in close contact with an infected person.^2^ Contact tracing was used as early as in the beginning of the 20^th^ century, in Scotland to contain sexually transmitted diseases (STDs).^3^ This strategy is commonly used for HIV, tuberculosis, and Ebola virus disease.^4,5^

Health inequities correspond to differences in health (mortality, morbidity) systematically linked, for example, to gender, socio-professional categories, or geographic areas. They are distributed according to a “social gradient of health”, where “each social class has a higher level of mortality and morbidity than the class immediately above”.^6^ The presence of health inequities can be explained by social, economic, political, and cultural determinants. Systematic and avoidable, they constitute a facet of social inequalities. In an epidemic context, health inequities are often exacerbated. For example, during 2009 H1N1 influenza pandemic, the mortality rate was higher within the most deprived population in England than the rest of the population, and higher in urban areas than rural areas.^7^ The Ebola virus disease epidemic in 2013 also affected more women than men due to their traditional role as caregivers.^8^ Nowadays, the current COVID-19 pandemic is exacerbating inequalities in incidence and mortality rate, according to ethnicity, socio-economic status, or living areas.^9^

Considerations related to the planning of public health interventions, and particularly those related to addressing health inequities, are paramount. For example, health inequities can increase within a population if the principle of “proportionate universalism”, whereby the intensity of effort is tailored to the needs of populations and their health status, is not integrated into the design of public health interventions. For example, containment and self-isolation increased food insecurity among the poorest during COVID-19 in the UK.^9^ Concerns have also emerged regarding contact tracing interventions, especially those using digital tools, such as smartphone applications. Indeed, these interventions could exacerbate health inequities by not reaching some geographical areas and populations due to a lack of internet access and smartphones.^10^ Studies have already shown the importance of examining how these interventions are designed to improve health policies.^11^ Some tools, such as the REFLEX-ISS tool, have also been created to raise awareness among stakeholders of how to incorporate health inequities when planning an intervention.^12^

The aim of this review was to identify if and how contact tracing interventions, in the context of outbreaks and epidemics, have considered health inequities.

**Contact tracing during COVID-19 pandemic worldwide**

South Korea rapidly implemented meticulous contact tracing, which led to the early identification of clusters.^13^ In addition to manual contact tracing, authorities have used records from credit card transaction history or information on their GPS location.^14^ In Japan, local governments played a large role in contact tracing investigations by reacting earlier than the national government authorities.^15^ Several contact interventions have been based on regional or district authorities (Finland, Spain, Italy, the Netherlands, Canada) whereas other countries have used a more centralized approach for contact tracing (Portugal, Russia, Luxembourg).^16^ Some countries have encountered difficulties related to trust in the government. For example, in North-Carolina (USA), 48 % of COVID-19 cases did not report their contacts in July 2020.^17^

Mobile apps for contact tracing have had varied success in different countries. In South Korea, China and Singapore, the adoption of mobile apps has been touted as key in containing the pandemic, due to a high level of uptake from the population. However, in Europe and in North America privacy issues and the lack of acceptability have undermined the use of such digital tools.^18^ The Finnish application, which uses Bluetooth technology, was downloaded by about 42 % of the Finnish population ^16^ whereas in France, « Stopcovid » apps using Bluetooth technology was download by 3% of French population.^19^ Finally, some studies have alerted that contact tracing apps can further increase health inequities by excluding the elderly or people who do not have the means to have a smart phone.^20^

## Method

We chose to conduct a rapid review of the literature as it allowed us to synthesize, with rigour and in a relatively short period of time, the state of knowledge on a specific research question. The rapid review was preferred to a full systematic review as our goal is to provide rapid information for public decision-makers, stakeholders and researchers.^21,22^

The method of analysis and the inclusion and exclusion criteria for this rapid review are detailed in an online protocol.^23^ The synthesis of the articles followed the recommendations of the PRISMA extension for scoping review (PRISMA-ScR) method.^24^

### Research strategy

The research strategy was developed in collaboration with librarians from the Research Institute for Development and the University of Montreal. We conducted our searches in the electronic databases MEDLINE and Web of sciences. The following keywords were used to define our queries (Appendix 1): “contact tracing”; “design*”, “plan*”; “disease*”, “epidemic*”, “pandemic*”. The references were exported and processed with the Automated Classifier of Texts on Scientific Studies (ATCER)^25,26^ tool to assess their degree of empiricism. Thus, the ATCER tool distinguishes empirical studies, based on qualitative, quantitative and mixed methods, from editorials, literature reviews or professional guidelines.

To be included in this rapid review, articles had to include a description of the design or concept of a contact tracing intervention in the context of an outbreak or epidemic. Our research focused on articles published in peer-reviewed scientific journals. We selected recent articles published between 2013 and July 2020. We selected 2013 as the beginning of our search in order to identify articles related to the Ebola outbreak in West Africa. Publications were included if they were written in English, French, Spanish, Portuguese or Chinese.

As we only wanted to retain scientific articles based on empirical data and avoid theoretical or methodological scientific articles, we used the ATCER tool to assess the empiricism of each article, and a threshold of 50% was used (which is the cut-off of ATCER to determine if an article is empirical or not). Grey literature and pre-publications were excluded from our research. We conducted our initial search between July 22 and July 31, 2020, with an update conducted on November 11, 2020.

### Selection of studies

All identified studies were exported into Rayyan QCRI software. Two reviewers (IM, LT) first independently assessed the relevance of the titles and abstracts on the basis of the defined inclusion and exclusion criteria. In case of disagreement between these two reviewers, a third reviewer (KO) decided. These two reviewers then independently assessed the full text relevance of the previously selected articles. Any disagreement involved the third reviewer.

### Study Characteristics, Quality Assessment, and Data Extraction

The dimensions studied to determine whether or not health inequities were considered in the design of contact tracing interventions in articles were: i) the inclusion of health inequities in the rationale for the intervention, ii) the populations targeted by the intervention, iii) the design of contact tracing, and iv) the presence of health inequities in recommendations for improving design of future interventions.

A first reviewer extracted data from the selected articles and a second reviewer verified the extracted data. The information extracted included information on the article (authors, title, year, country, type of publication, and type of evaluation if applicable), information on contact tracing and equity (level of jurisdiction, diseases, description of contact tracing intervention, participants, reference to equity, and main objectives), the Mixed Methods Assessment Tool (MMAT)^27^ grid to describe the methods used, the results of the study (outcome, methodological limitations and main conclusions), and the Template for Intervention Description and Replication - Population Health and Policy (TIDieR-PHP)^28^ grid to describe the content of the interventions.

## Result

### Description of the studies

We identified 230 references and 39 met the inclusion criteria (Figure 1). The analysis of the full texts led to the final selection of 7 relevant references^29,30,31,32,33,34,35^ (Annex 1).

**Figure 1:**
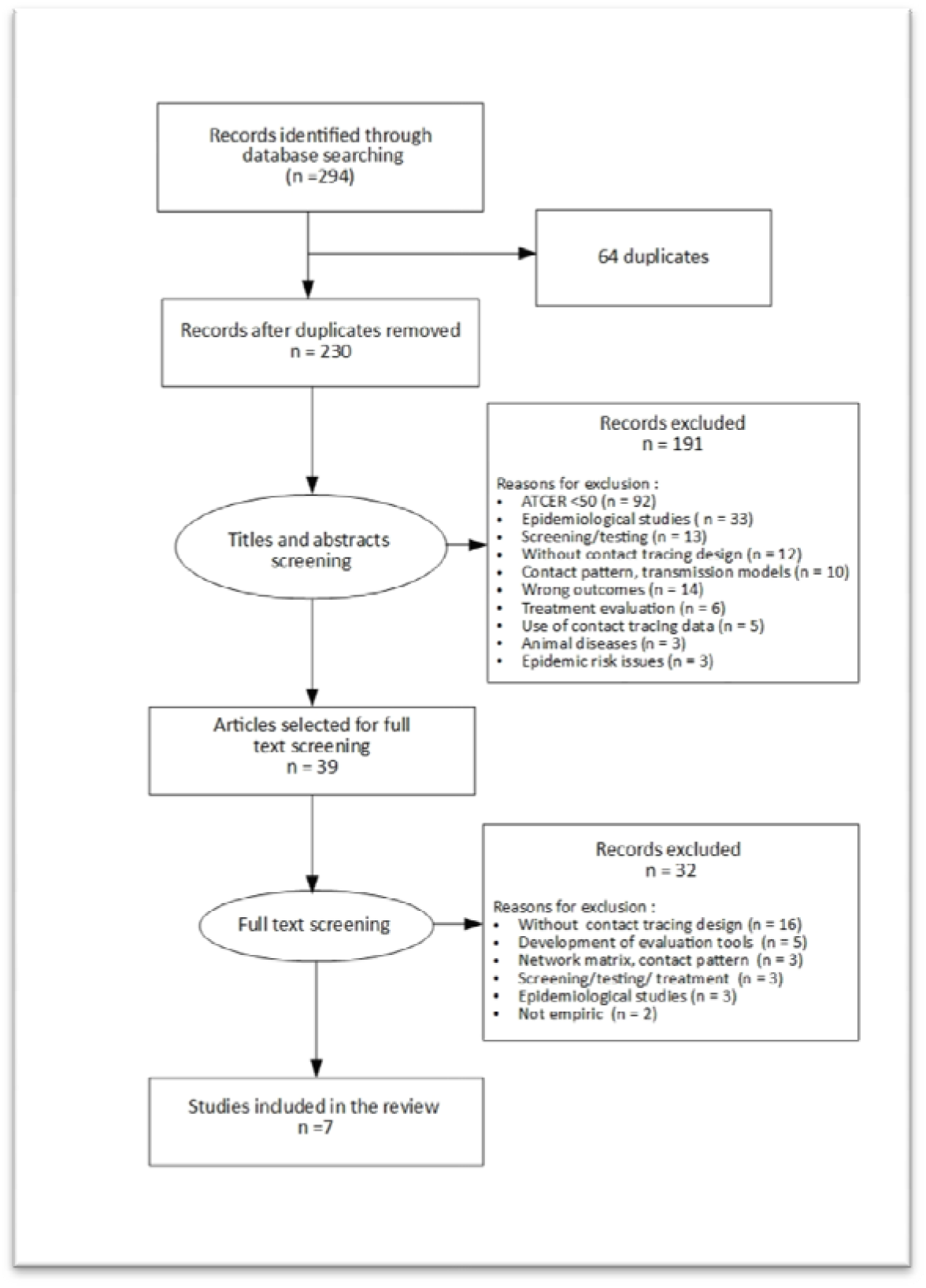
PRISMA diagram

These articles were based on contact tracing interventions conducted in Africa (n=5) and Asia (n=2). The contact tracing interventions include Ebola (n=2), tuberculosis (n=2), Lassa fever (n=1), COVID-19 (n=1), and HIV (n=1). These articles included experimental designs (n=4) and observational designs (n=3), with most of the studies measuring effectiveness of the intervention (n=5) with two studies being descriptive in nature.

The selected articles differed in the tools used for the deployment of contact tracing interventions. Three interventions prioritized the use of technological tools in contact tracing, such as smartphones equipped with an application^29,30,31^ or real-time location systems.^32^ Three interventions planned contact tracing based on telephone tracking and manual contact case entry.^33,34,35^

### The inclusion of health inequities in the rationale for the intervention

For interventions using contact tracing applications and real-time location, the rationale for this mode of contact tracing was the speed of data collection and the tools’ reliability^30,31,32^, low cost,^32^ and better accessibility to cases in isolated areas.^29^ For interventions based on manual data collection and telephone follow-up, the rationale for these approaches were the ease of implementation in an epidemic context^34^ and their ease of implementation during household contact tracing. ^33^ In Kenya, Community Health Volunteers (CHVs) were chosen to perform contact tracing by phone and manually as they provided links between the community and the health system, which can improve healthcare access and community empowerment.^35^ None of the articles mentioned incorporating health inequities into their intervention.

#### The populations targeted by the intervention

The populations studied were direct contacts of laboratory-confirmed cases, whether they were household contacts or by occupation (healthcare workers).^29,30,31,32,33,34,35^ No intervention included a sub-group of the population, whether in terms of income, socio-professional category, gender, or geographic area.

#### The design of contact tracing

Contact tracing interventions involve following up suspect or probable cases. For the two studies that occurred within the context of the West African Ebola epidemic and one for Lassa fever in West Africa, the follow-up took place by an epidemic control team.^29,30,34^ Daily temperature and monitoring of clinical signs and symptoms were recorded over a period of 21 days for all suspect cases. For the HIV intervention in Kenya,^31^ individuals were contacted one to three times by telephone. If the person was unreachable, the contact tracing team visited the contact person directly (one to two visits) to convince him or her to be tested. For the tuberculosis intervention in Vietnam,^33^ contact tracing within the household was conducted three times every six months in the clinic. A questionnaire on symptoms was administered and included a physical examination and x-rays. In order to encourage as many contact cases as possible to visit the clinics, contact cases within the household received a compensation of one dollar for the journey. Finally, for tuberculosis intervention in Kenya, contact tracing in the household of a confirmed case was conducted by CHVs. Household contacts of tuberculosis cases were screened by CHVs by phone or through household visits and ensured that positive cases arrived at the health facility for healthcare.^35^ None of the interventions adapted their follow-up to the income, gender, or the socio-professional profile of the target population.

#### The presence of health inequities in recommendations for improving future interventions

A few of the articles provided recommendations for improving contact tracing interventions. These recommendations mainly concerned mobile contact tracing applications and the technical problems encountered during the intervention (e.g., problems related to internet access, team training, lack of computer equipment, lack of technical support).^29^ No recommendations were provided on how to include health inequities, for example, adapting the intervention according to income, geography, or gender. One article that involved manual contact tracing for HIV in Kenya recommended focusing on the regions and populations most at-risk, promoting access to antiretroviral therapy.^31^ Another article that involved contact tracing for tuberculosis in Kenya with CHVs, recommended targeting the informal labour sector to increase contact tracing and screening among men, often absent during CHVs visits.^35^ However, there was no specific reference in the article about health inequities.

**Health inequities and contact tracing interventions**

- None of the studies mentioned health inequities nor included a dimension of health inequities.
- No intervention targeted a sub-group of the population, for example, on the basis of income, profession, gender, or geographic area.
- None of the studies adapted their evaluation to include measures of income, gender, or profession of the participants.

## Discussion

This rapid review demonstrated that health inequities have not been accounted for with contact tracing interventions that were included in this review. No specific adaptations were made in the planning or implementation of contact tracing interventions to consider the particular needs of certain sub-groups of intervention population.

None of studies included in the review proposed actions proportionate to target sub-groups when planning the contact tracing intervention. This principle could be incorporated in contact tracing interventions by targeting different sub-groups and disadvantaged groups, depending on social determinants, such as age, socioeconomic status, profession, ethnicity, or geographic area. For example, communication tools during contact tracing interventions could be adapted to low literate population (by including pictographs), migrants who do not speak official languages (by including other languages), or to disabled people (audio messages).

Tools exist that can facilitate the inclusion of health inequities in the design of public health interventions. For example, the REFLEX-ISS tool is for decision-makers, researchers, and stakeholders, which helps to analyze, the consideration of health inequities during an intervention and also supports dialogue between stakeholders throughout the life cycle of the intervention (from planning to sustainability of the project).^12^ A creation of a guide, bringing together the evidence of how health inequities should be considered throughout the intervention process, from the design to evaluation would be an invaluable tool to practitioners, researchers, and decision makers. This is particularly important as planning is underway with the rollout of COVID-19 vaccines. Furthermore, in the TIDieR-PHP reporting guidelines, there are no categories directly related to health inequities. This addition in the guidelines would encourage researchers and public practitioners to consider aspects of health inequities when planning and implementing public health interventions.

### Limitations of the study

Relevant articles could have been excluded from the review, despite the screening having been performed by two reviewers. In addition, studies may have been excluded that did account for inequities in their contact tracing intervention but was not specifically mentioned in the article.

**Recommendations for improving the consideration of health inequities**

1. Apply proportionate universalism rather than targeting a specific population
2. Use health inequities reflection tool(s) when designing public health interventions such as REFLEX-ISS
3. Create a guide as a resource that describes how health inequities should be considered in public health interventions, from design to evaluation
4. Integrate categories of inequity within the TIDieR-PHP framework

## Conclusion

This rapid review demonstrates that health inequities were not included in several contact tracing interventions that were conducted in outbreak or epidemic settings. The emergence of COVID-19 has prompted governments to act swiftly, including the implementation of contact tracing interventions. However, these interventions can increase health inequities between different population sub-groups. It is imperative that practitioners, researchers, and decision makers take health inequities into account when designing contact tracing interventions in outbreak or epidemic settings.

## Supporting information

PRISMA checklist

## Data Availability

Not applicable

## Acknowledgements

The authors would like to thank Laurence Goury, librarian at the IRD, and Julie Desnoyers, librarian at the University of Montreal, for their remarks and advice on the definition of the search strategy and the queries on the bibliographic databases. The authors are also grateful to Anne Guichard, associate professor at Laval University, for her comments on the manuscript.

## Conflict of interest

The authors declare that they have no known competing financial interests or personal relationships that could have appeared to influence the work reported in this paper.

## Funding sources

The research project is funded by the French National Research Agency (ANR) (ANR-20-COVI-000) and the Canadian Institutes of Health Research (202002OV7-440254-CV2-CFCA-120159).

## Ethical Approval

Not applicable.

## Appendix

**Annex 1:** MEDLINE and Web of Science queries (filled in on 29/07/2020 and on 11/11/2020)

| **Database** | **Keywords** | **Number of**<br>**references**<br>**listed**<br>**(07/2020)** | **Number of**<br>**references**<br>**listed**<br>**(11/2020)** |
| --- | --- | --- | --- |
| MEDLINE | ((contact tracing[MeSH]) OR ("contact tracing"[Title/Abstract])) AND<br>(design*[Title/Abstract] OR plan*[Title/Abstract])<br>AND (( (disease*[Title/Abstract] OR<br>epidemic*[Title/Abstract] OR<br>pandemic*[Title/Abstract])) Filters: Full text,<br>Chinese, English, French, Portuguese, Spanish,<br>MEDLINE, from 2013 - 2020 | 130 | 38 |
| Web of<br>Science | (((((ts ="contact tracing") AND ((ts =design*) OR<br>(ts=plan*) ) AND ((ts=disease*) OR<br>(ts=pandemic*) OR (ts=epidemic*) ))))) AND<br>LANGUAGE: (English OR Chinese OR French<br>OR Portuguese OR Spanish)<br>*Indexes=SCI-EXPANDED, SSCI, A&HCI, CPCI-S,*<br>*CPCI-SSH, ESCI Timespan=2013-2020* | 74 | 52 |

## Notes

### Competing Interest Statement

The authors have declared no competing interest.

